# Structured Error Analysis and Corrective Actions in Clinical Laboratory Practice: An Analysis of 7226 External Quality Assurance Participations

**DOI:** 10.64898/2026.04.02.26350023

**Authors:** Bernhard Strasser, Sebastian Mustafa, Matej Holly, Monika Grünberger, Anita Stelzhammer

**Author notes:** **Corresponding Author:** Bernhard Strasser, Department/Institution: Institute of Clinical Chemistry and Laboratory Medicine, Klinikum Wels-Grieskirchen, Postal Address: Grieskirchnerstraße 42, 4600 Wels, Austria.

## Abstract

**Background:** External Quality Assurance (EQA) is an essential component of modern laboratory medicine. Current scientific evidence on EQA focuses primarily on the analyses carried out by EQA providers while relatively little research has been conducted in individual clinical laboratories.

**Methods:** In this retrospective single-center observational study in a clinical laboratory, EQA results were analyzed over a period of four years (2021–2024). The evaluation was based on EQA action reports documented in the institute’s internal quality management system. Deviations were classified according to department, type of discrepancy, root cause category (analytical, preanalytical, systemic, unidentifiable), and measures taken.

**Results:** A total of 7226 EQA participations were evaluated during the observation period. The overall error rate remained consistently low, ranging between 0.8% and 1.6%, with no significant change over time (p = 0.87). Most deviations occurred in the departments of clinical chemistry and immuno/autoimmune diagnostics (p < 0.001). These were predominantly quantitative discrepancies (false low/false negative or false high/false positive). Root cause analysis showed a clear dominance of analytical causes (p < 0.001), while preanalytical and systemic causes were identified less frequently. In most cases, corrective measures, such as re-analyses, recalibrations, process adjustments, or staff training, were implemented promptly. Hard structural measures, such as changing methods or discontinuing tests, were rarely necessary.

**Conclusion:** In a clinical laboratory, EQA is an important tool for structured error analysis and continuous quality improvement. Consistent processing of deviating EQA results goes hand in hand with stable analytical performance and a low error rate.

## Background

External quality assurance (EQA) is now considered an indispensable constant in clinical laboratories. EQA is a relevant part of quality control in clinical laboratories, and the use of such initiatives is also supported by international standards such as ISO 15189:2022 and ISO/IEC 17043:2023. As there is no international consensus on the legal framework, EQA is mandatory in some countries but not in others to the same extent. A key dimension of EQA is to ensure the accuracy and consistency of diagnostic procedures through standardized performance evaluations, thereby prioritizing patient safety (1, 2). In addition to ensuring and testing diagnostic quality, EQA can be utilized to make further and continuous improvements. In the context of harmonizing laboratory results, EQA programs can provide information on whether results from different measurement systems are comparable. Thus, EQA can help identify systematic differences between clinical laboratory systems and provide impetus for promoting standardization between different clinical measurement methods (1, 3-5). A further potential of EQA is its facilitation of trend analysis. EQA programs allow longitudinal assessments of test performance for different parameters, participants, or clinical laboratories (6, 7). EQA not only allows methodological data to be collected and collectively analyzed but also enables methodological developments to be tracked over the years (1). Using more modern approaches, EQA programs have also been linked to cloud-based learning units, resulting in significant improvements in diagnostic performance (8). A survey showed that users (clinical laboratories) saw EQA’s comparative feedback, ranking information, improved patient safety, error reduction, and improved internal decision-making, as perceived benefits. Difficulties in data collection, lack of knowledge about the EQA program, lack of suitable laboratory information system (LIS) indices, lack of time, and lack of organizational guidelines were cited as factors that made participation in EQA difficult (9). The basis for continuous learning is a well-founded evaluation measure applied by the EQA provider, whose report is based on target values, acceptable performance limits, and evaluation of the participants’ results. Most reports also contain expert comments with references to possible causes of errors and clinical or analytical recommendations (10). Especially in cases of high methodological variation between laboratories, consensus assessments within the peer group are preferred (11). In addition, it is necessary to identify a robust method for evaluating analytical performance in EQA contexts (12). A challenge for EQA providers is the varying participation in EQA programs, which is due to differing legal frameworks varying greatly across individual countries. And since accreditation is not mandatory in most countries, there is no uniform pressure to participate in EQA, which leads to difficulties in harmonizing quality standards (13).

## Objective

Currently, numerous EQA publications reflect the perspective of providers. These are undoubtedly valuable scientific contributions, providing important information on overarching interindividual laboratory medicine content (e.g. standardization, harmonization). They also enable cross-sector information to be collected (provider- or company-specific differences, clinical vs. non-clinical laboratories) and problem areas to be identified. (14, 15) In contrast, this study aims to shed light on the perspective of the user, examining the measures a clinical laboratory implements in the event of deviating EQA results.

## Methods

### Study Procedure

In this retrospective analysis, data from 2021−2024 was evaluated by the Institute of Clinical Chemistry and Laboratory Medicine, Wels-Grieskirchen Hospital, Wels, Austria. The data comprised reports used for the analysis are archived in the local institute’s internal quality management system. These reports provide a clear overview of the measures taken in response to abnormalities identified during EQA. The reports follow a standardized structure, containing organizational information, such as information granularity (i.e., who was the primary creator, who approved it, and when were all employees informed), document type, and a sequential EQA measure report number. The reports also include more EQA-specific data, such as a description of the EQA, the parameter affected and the deviation, the laboratory area affected, a cause analysis, an assessment of the extent of the error, any immediate and corrective measures, and accompanying documents (e.g., confirmation of participation, a confirmation certificate, and longitudinal assessment). During data collection, the information contained in the problem reports was systematically extracted and converted into a uniform analysis format. First, the year of the incident was entered into the template, followed by the affected department within the laboratory institute (e.g., Hematology, Hemostasis, Immuno/Autoimmunology, Clinical Chemistry, Toxicology/TDM, Blood Bank, or Genetics). The type of deviation was recorded according to the metrological categories used in the quality management system: false low/false negative, false high/false positive, borderline deviation, or precision-related deviation. The documented root cause analysis was also systematically categorized into analytical, preanalytical, systemic, or unidentifiable causes. Analytical causes were related to methodological aspects. In particular, device-related problems, such as calibration, reagents, test system characteristics, or measurement method limitations, were included. Pre-analytical causes covered all influencing factors that could occur prior to analysis. Accordingly, sample collection, transport, storage, or handling-related management of interlaboratory test material were included in this category. Systemic causes included all external or structural influencing factors that were outside the immediate scope of the analysis. These included EQA-specific features, cross-method variability, EQA specifications, EQA provider target values, organizational processes, and software frameworks (e.g., LIS configurations or interfaces). The category “unidentifiable” was used in the root cause analysis if the action report did not allow a clear assignment. Retesting and correction submission were subsumed under corrective action. In addition, corrective action included hard measures such as process correction, device or test kit replacement, or establishment of a new method. Checklists and dual control principle were summarized under preventive action, and continuing internal education under training. The measure “communication” refers to a team discussion, re-evaluation, and manufacturer request. “No action” was selected if reference was made to external follow-up. In addition, contextual information was evaluated qualitatively using a narrative synthesis and specified methodological features, sample problems, or round-robin test-specific influencing factors. Figure 1 provides an overview of the study procedure.

**Figure 1:**
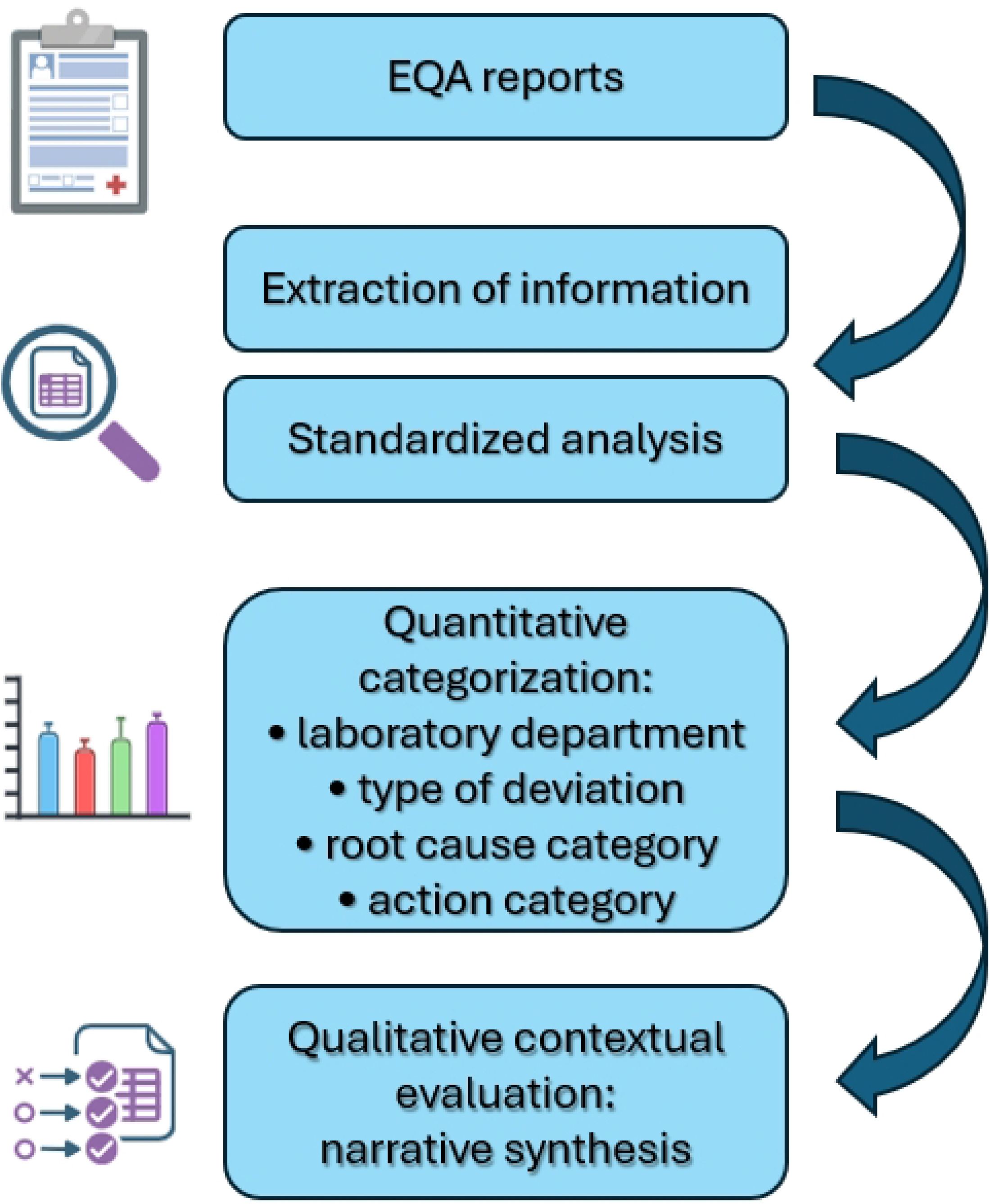
The flowchart illustrates the study procedure.

In this retrospective study, no direct patient data was processed, rather data from quality management related to EQA. Therefore, publication of the data is in accordance with the General Data Protection Regulation. The Institutional Review Board of the State of Upper Austria, Johannes Kepler University, deemed this study exempt from review.

### Statistics

The data analysis included both descriptive and inferential statistical methods. Absolute and relative frequencies were determined for all categories. In the case of multiple entries, particularly in the category of measures, these were systematically separated into individual measures and consequently included in the evaluation as independent units. Chi-square tests were performed to check whether the observed frequency distributions of the categorical variables (e.g., discrepancy type, root cause category, or corrective action category) were uniformly distributed or differed from each other. Linear regression was performed to analyze temporal trends in the annual error rate. A two-sided significance level of α = 0.05 was used. Statistical analyses were performed using the software “Analyse IT – Medical Edition”.

### Results

Between 2021 and 2024, a total of 7226 EQA were conducted at the Institute for Medical and Chemical Laboratory Diagnostics at the Hospital Wels-Grieskirchen, as shown in Table 1. The error rate remained below 2% over the period (Figure 2) but showed annual fluctuations. Accordingly, the absolute number of failed EQAs ranged between 15 and 29 (0.8 – 1.6%) per year. A linear trend analysis revealed no statistically significant change in the error rate over the observation period (p = 0.87), indicating an overall stable and consistent performance quality.

**Table 1:** Annual number of External Quality Assurance (EQA) participations, failed and passed EQAs, and corresponding error rates from 2021 to 2024.

**Figure 2:**
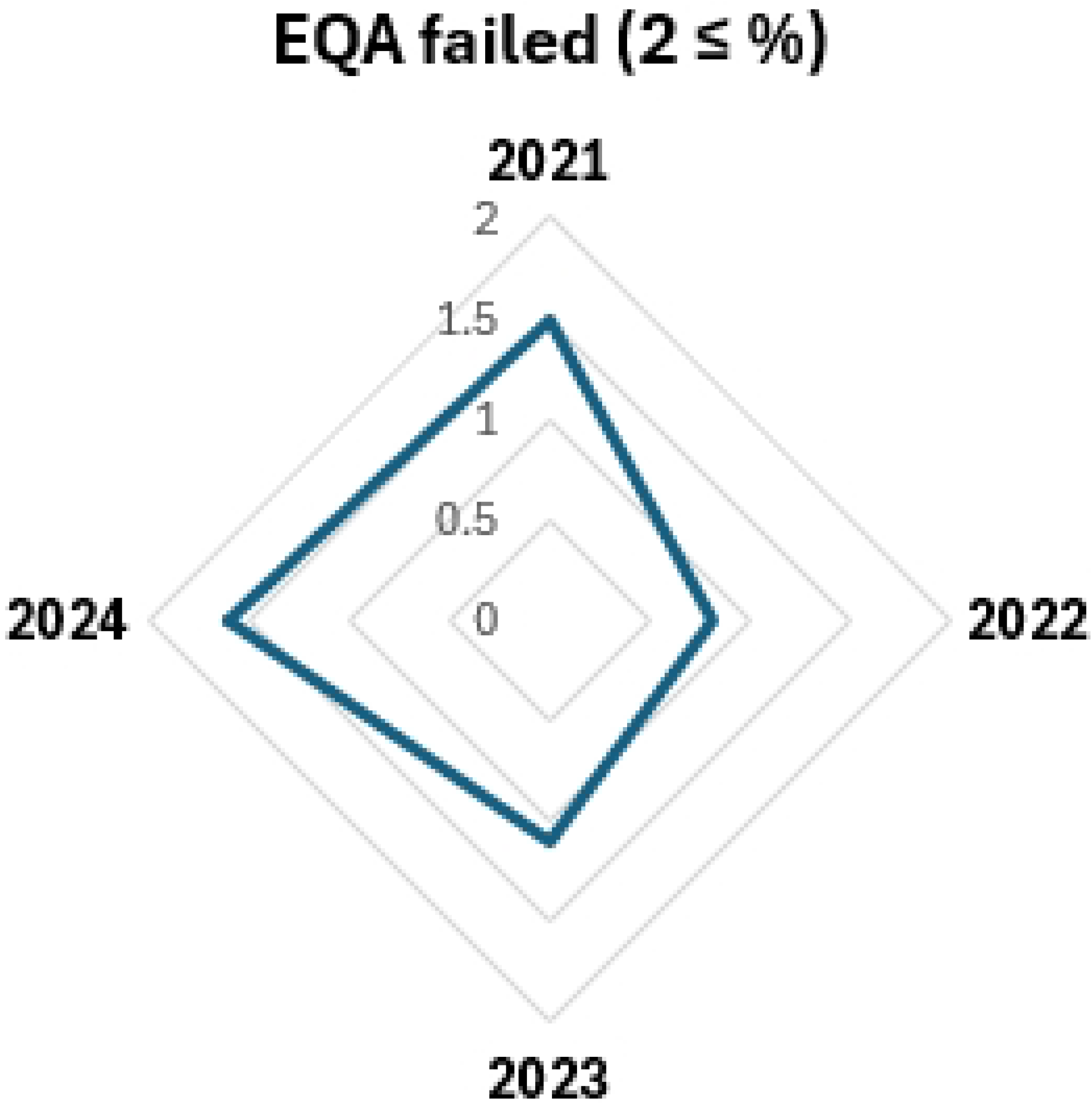
Annual External Quality Assurance (EQA) error rate from 2021 to 2024.

Most deviations occurred in clinical chemistry (n = 34, 37.0%), followed by immuno/autoimmunology (n = 28, 30.4%). Together, these two areas accounted for more than two-thirds of all deviations (67.4%). Hematology contributed 14.1% (n = 13), while smaller proportions were observed for toxicology/TDM (n = 6, 6.5%), hemostasis (n = 5, 5.4%), and genetics (n = 5, 5.4%). The lowest number of deviations was observed in the blood bank (n = 1, 1.1%). The distribution of deviations across laboratory disciplines was uneven, indicating a significant imbalance between departments (p < 0.001). The evaluation of the discrepancy type showed that the most common categories were false low/negative (n = 33) and false high/positive (n = 30), whilst no evaluation possible was the least common (n = 7). Most of the relevant deviations originated from quantitative discrepancies rather than categorical or non-assessable ones. The frequency distribution of the affected departments and the discrepancy types are shown in Figure 2.

The analysis of the root cause categories showed a dominance of analytical causes, accounting for most of the identified deviations (p < 0.001). The relative frequency distribution shows that most deviations were managed through corrective measures, whereas preventive measures were implemented less frequently (p < 0.01). Figure 3 shows the frequency distribution of the root cause category and the measures implemented.

**Figure 3:**
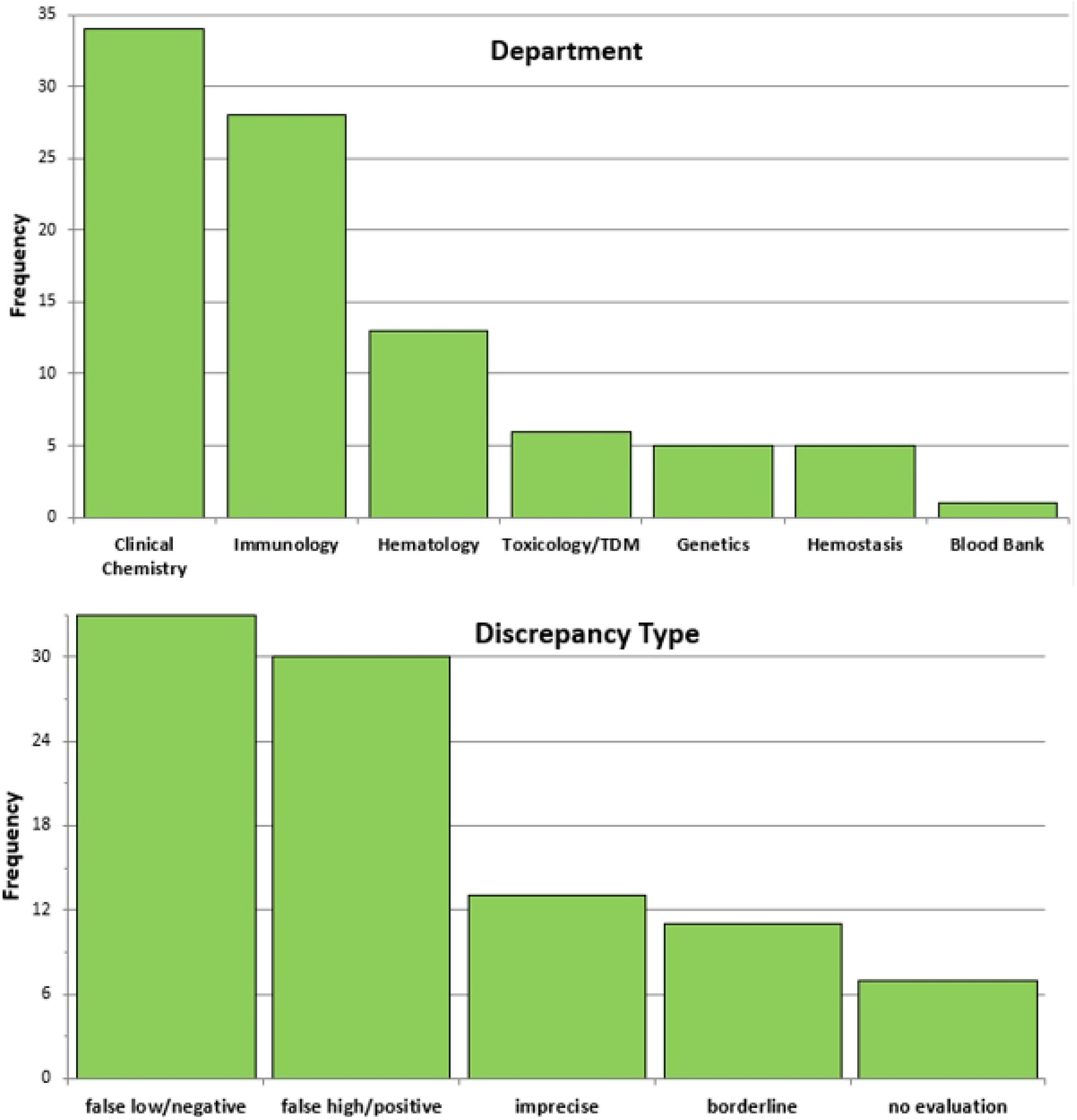
Distribution of deviating External Quality Assurance (EQA) results by laboratory department and type of discrepancy.

**Figure 4:**
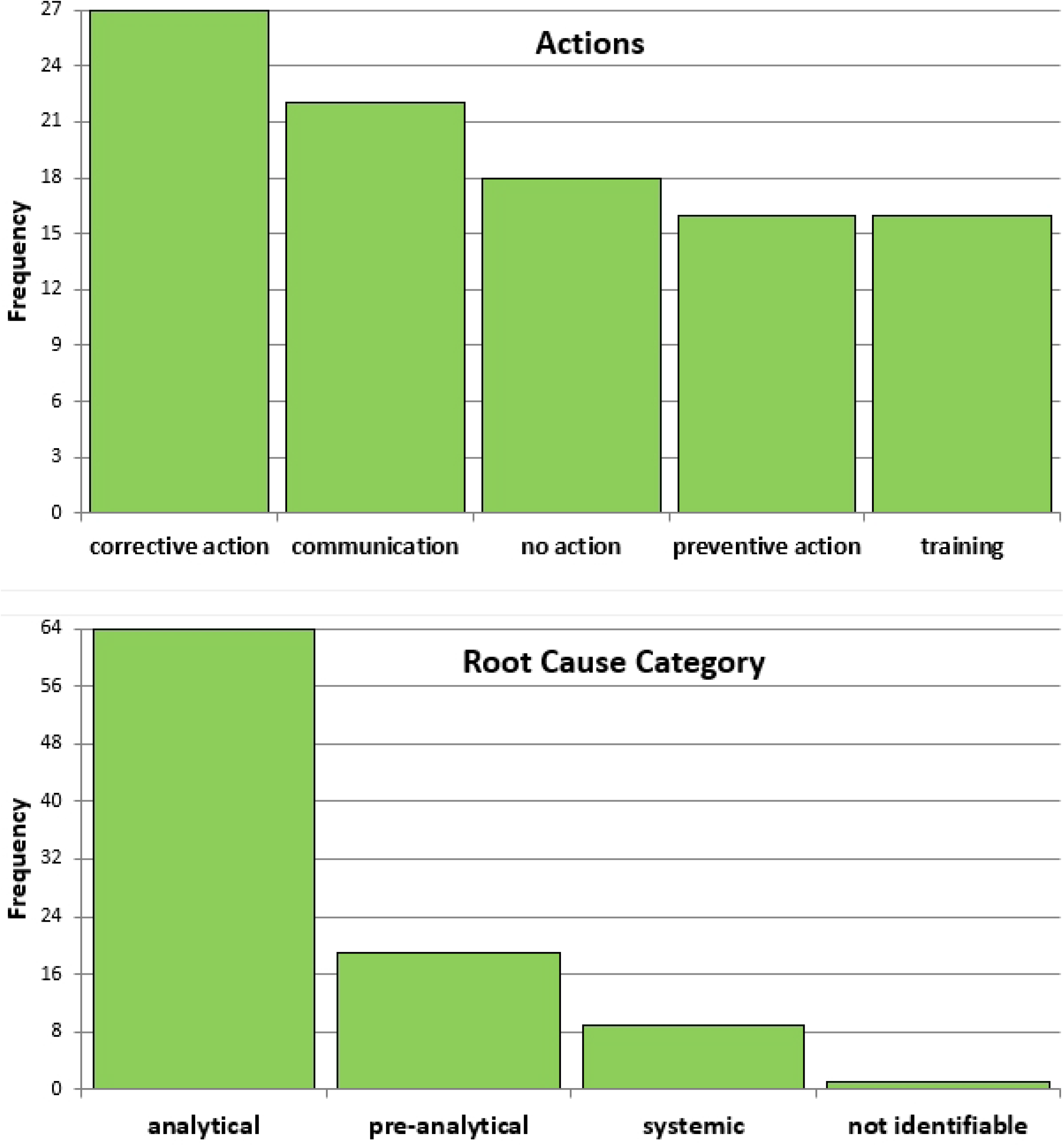
Frequency distribution of root cause categories and implemented measures following deviating External Quality Assurance (EQA) results.

For qualitative evaluation, a narrative synthesis was performed in which the contents were assigned to the thematic categories listed in Table 2. These categories comprised both cause-related and action-related aspects.

**Table 2:** Categories derived from narrative synthesis of deviating External Quality Assurance (EQA) reports, including cause-related and action-related aspects.

The predominant causes of the EQA discrepancies were not due to systemic errors, but the most frequently described issues were method- or test system-related characteristics, such as calibration deviations, matrix-related effects, or known limitations of specific assays. Several deviations were preanalytical in nature (e.g., sample instability, incorrect reconstitution, or transport problems). Another group was due to administrative causes, such as data or unit entry errors, incorrect method assignments, or upload problems at the EQA provider. In addition, cases were documented in which results were measured correctly but inadequately interpreted or classified. In these cases, specific training measures were then always taken. A smaller group of discrepancies was attributable to external factors on the part of the EQA provider (e.g., insufficient target values, delayed delivery, or method-specific evaluation problems). Genuine analytical errors did occur but were almost always successfully resolved through recalibration or process corrections. Further details can be found in the Supplementary Material.

Most deviating EQA results were managed by corrective measures and a small subset required hard measures such as major structural interventions. These included method changes, assay discontinuations with subsequent outsourcing of analyses, or restrictions in routine reporting as shown in Table 3.

**Table 3:** Hard measures following External Quality Assurance (EQA) results. Abbreviations: aPS/PT, anti-phosphatidylserine/prothrombin antibodies; VWF:RCo, von Willebrand factor ristocetin cofactor assay; VWF:CB, von Willebrand factor collagen-binding assay; EDDP, 2-ethylidene-1,5-dimethyl-3,3-diphenylpyrrolidine; IgA, immunoglobulin A; IgG, immunoglobulin G; CSF, cerebrospinal fluid; NGS, next-generation sequencing; HBA/HBB, hemoglobin subunit alpha/beta genes; FACS, fluorescence-activated cell sorting; NK, natural killer; CNV, copy number variation; *CALR*, calreticulin gene.

## Discussion

Within the framework of EQA, it is important to highlight the role of clinical laboratories and the potential for learning. Kristensen et al. provide a structured approach to troubleshooting deviating EQA results. (16) First, it is necessary to determine what type of error has occurred, making a rough distinction between internal laboratory errors (the most common cause), errors on the part of the EQA provider, and external errors caused by manufacturers (e.g., reagents). Errors can occur as transcription errors, preanalytical during sample transport or sample storage or preparation as well as during measurement, evaluation, or interpretation (16). In the present study, a comparable categorical root cause classification was made. The most frequent deviations were related to classical analytical errors, which were always responded to with appropriate measures. In addition, this study also dealt with systematic deviations. These errors were not method-specific, but concerned overarching processes, infrastructure, or IT and organizational structures.

From a user perspective, it is necessary to integrate the EQA reports into the internal quality management system. Furthermore, measures should be taken in the event of inadequate performance. Root cause analyses can reveal operating errors, problems with sample handling, calibration, reagent batch changes, or equipment problems (10). This study demonstrated that analytical errors are not primarily caused by gross errors but occur mostly as discrete deviations. However, in a clinical laboratory, it is not only analytical errors but also extra-analytical errors that must be taken into account; this applies to core laboratory processes as well as to participation in EQA programs. (17, 18) Many of the preanalytical deviations can be compensated for by routine use of improved supportive processes (LIS, digitization) or faster processing. But, EQA is not just a monitoring tool; it is a learning tool. This study demonstrated that in most cases, corrective or communicative measures were initiated immediately, indicating an established quality management system. Staff training was a recurring component of the measures initiated. Most of the deviations identified were classified as clinically and/or analytically irrelevant, which means that overall patient safety can be rated as high. In individual cases, hard measures were taken and EQA discrepancies led to structural measures, such as a discontinuation of individual analyses or switching to alternative analytical methods. However, such measures remained rare and mainly affected tests with clearly identified methodological limitations. In most cases, deviations could be corrected through corrective or organizational measures without requiring a permanent change in the range of methods used.

Based on the analysis of 7226 EQA participations, this study shows that the clinical laboratory under investigation has stable and consistent performance. The error rate remained below 2% throughout the observation period and showed no significant change over time. Comparable data showed that the error rate in central medical laboratories participating in EQA ranges between 1% and 2%, and that as the number of tests performed in a medical laboratory increases, so does its performance in EQA, since greater experience and improved routine generally contribute to more stable processes. (19) Analysis of the deviations in our study showed that discrepancies were predominantly analytical in nature and manifested primarily as quantitative deviations, while systemic or preanalytical causes played a minor role. Deviations did not occur evenly across all laboratory departments but were concentrated in clinical chemistry and immuno/autoimmune diagnostics. This observation may be due to the fact that inter-assay variability exists even in most biomarkers analyzed by immunoassays. (20, 21) There is a wide variety of methods, particularly in the field of autoimmune diagnostics. (1)This heterogeneity also increases the likelihood of method-related differences in EQA rounds. (22)

Nevertheless, this study has several limitations. Due to its retrospective design, the analyses are based on the quality and completeness of the EQA reports documented in the quality management system. In addition, this is a single-center study, which may limit the transferability of the results to other laboratory structures. Patient-specific clinical outcomes could not be analyzed, as quality-related data were evaluated only.

## Conclusion

EQA functions not only as a control instrument but also as a pivotal part of a continuously developing and improving quality management system. This makes EQA a practical tool for sustainable development in clinical laboratories as learning organizations.

## Data Availability

All relevant data are within the manuscript and its Supporting Information files.

## Research ethics

The Local Institutional Review Board of Upper Austria /Johannes Kepler University Linz determined that the study (submission no. 1169/2025, decision dated 13 May 2025) is exempt from ethics review.

## Informed consent

Not applicable.

## Author contributions

BS wrote the first draft. All remaining authors commented on and revised the manuscript. All authors made substantial contributions to the manuscript. All authors approved the final version of the manuscript. All authors have accepted responsibility for the entire content of this manuscript and approved its submission.

## Use of Large Language Models, AI and Machine Learning Tools

None declared.

## Research funding

None declared.

## Data availability

The datasets generated or analyzed during the current study are included in the manuscript and/or the supplementary material.

